# Impact of mental disorders on active tuberculosis treatment outcomes: a systematic review and meta-analysis

**DOI:** 10.1101/2020.06.19.20135913

**Authors:** Ga Eun Lee, James Scuffell, Jerome T. Galea, Sanghyuk S. Shin, Elizabeth Magill, Ernesto Jaramillo, Annika C. Sweetland

## Abstract

**Background:** Comorbid mental disorders in patients with tuberculosis (TB) may exacerbate TB treatment outcomes. We systematically reviewed current evidence on the association between mental disorders and TB outcomes.

**Methods:** We searched eight databases for studies published from 1990-2018 that compared TB treatment outcomes among patients with and without mental disorders. We excluded studies that did not systematically assess mental disorders and studies limited to substance use. We extracted study and patient characteristics and effect measures and performed a meta-analysis using random-effects models to calculate summary odds ratios (OR) with 95% confidence intervals (CI).

**Findings:** Of 7,687 studies identified, ten were included in the systematic review and nine in the meta-analysis. Measurement of mental disorders and TB outcomes were heterogeneous across studies. The pooled association between mental disorders and any poor outcome, loss to follow-up, and non-adherence were OR 2.13 (95% CI: 0.85-5.37), 1.90 (0.33-10.91), and 1.60 (0.81-3.02), respectively. High statistical heterogeneity was present.

**Interpretation:** Our review suggests that mental disorders in TB patients increase the risk of poor TB outcomes, but pooled estimates were imprecise due to small number of eligible studies. Integration of psychological and TB services might improve TB outcomes and progress towards TB elimination.

## INTRODUCTION

To eliminate tuberculosis (TB) by 2030, the World Health Organization’s End TB Strategy calls for integrated, patient-centered care and prevention.^1^ This strategic pillar underscores the management of co-morbidities, including mental disorders, which have been identified in up to 70% of TB patients.^2^ TB and mental disorders share common risk factors including homelessness, substance use, and HIV infection which may affect health-related behaviours and treatment outcomes.^2^

A large body of work suggests an association between mental disorders and poor TB treatment outcomes,^2–9^ yet many studies were not primarily designed to answer this research question.^3–6^ Not all studies assess mental disorders systematically across all participants,^8,9^ making results difficult to interpret.^10^ This gap in evidence greatly impedes progress to formulate an evidence-based strategy to address mental health in TB patients.

The purpose of this study was to systematically review evidence on the impact of mental disorders on TB treatment outcomes. We only included studies that assessed mental disorders systematically for all TB patients. Our primary objective was to determine whether TB patients with concurrent mental disorders have poorer treatment outcomes than patients without mental disorders. We also examined evidence on delayed treatment initiation and medication adherence.

## METHODS

This systematic review and meta-analysis was conducted in accordance with PRISMA guidelines and the protocol was registered in the PROSPERO database (CRD42019122382). Ethical approval was not required for this study.

### Search strategy

We searched PubMed, Embase Classic, PsycINFO, Global Health, CINAHL, LILACS, Scopus, and Web of Science Core Collection databases for terms related to TB, mental disorder, and TB treatment outcomes. We used both free-text and subject heading (MeSH) terms adapted for use in each database. The MeSH terms search was: “Tuberculosis AND Mental disorders AND (Mortality OR Morbidity OR Death OR Prognosis OR Disease-Free Survival OR Treatment Outcome OR Drug Resistance OR Disease transmission, infectious OR Patient Acceptance of Health Care OR Treatment Failure OR Patient Compliance OR Lost to Follow-Up OR Patient Dropouts OR Treatment Refusal OR Health behavior OR Help-seeking Behavior OR recurrence)”. The full search strategy for PubMed is available in Table 1.

**Table 1.**
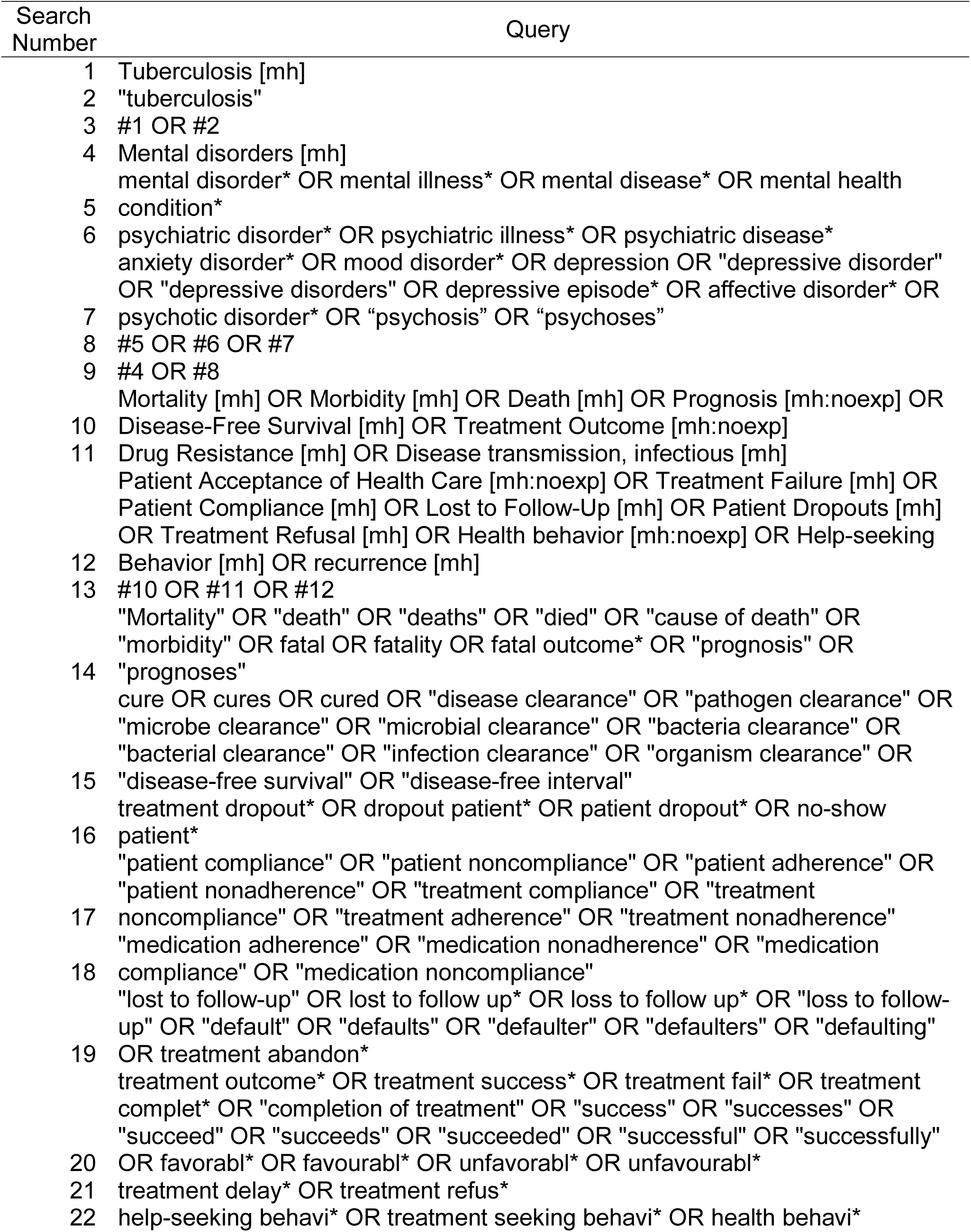
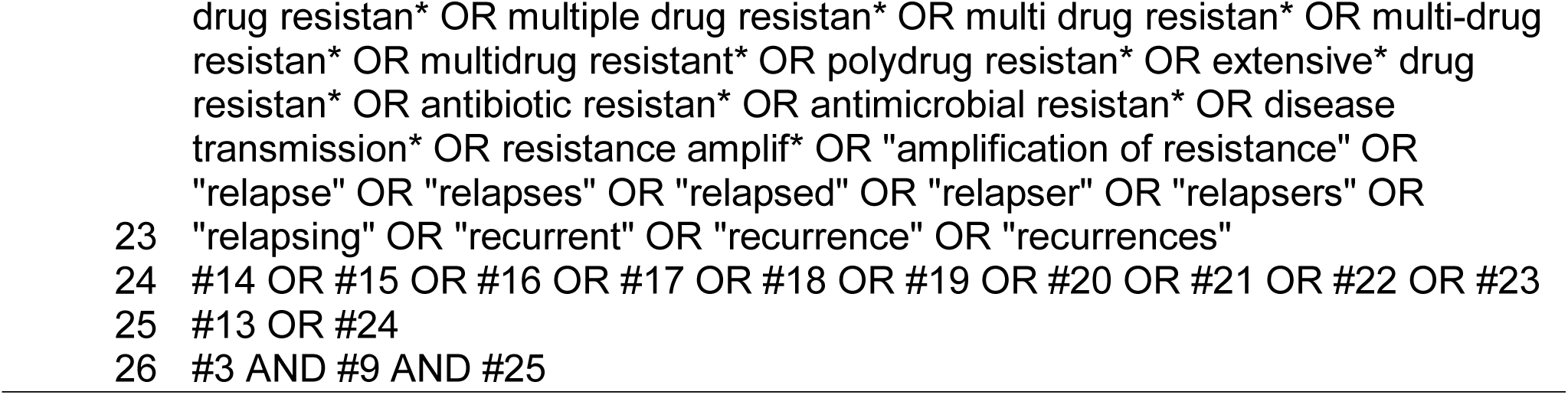
Search strategy for Pubmed

Database searches were conducted during October 2018. We included articles published between January 1, 1990 - towards the beginning of the Directly Observed Therapy, Short Course (DOTS) strategy - and October 29, 2018. At full-text screening, we reviewed articles published in languages familiar to the co-authors (English, French, Spanish, Portuguese, Korean). We also screened relevant citations in included articles and searched for subsequent publication of study protocols.

### Eligibility criteria

#### Study type and participants

Study types included cross-sectional, case-control, and retrospective and prospective cohort studies, as well as randomized controlled trials. We excluded qualitative studies, case reports and series, theses, literature reviews, meta-analyses, commentaries, editorials, conference abstracts, and latent TB studies. Studies included patients initiating or undergoing treatment for active TB in any setting, with or without multidrug-resistance (MDR).

#### Exposures and outcomes of interest

We defined mental disorders as mood disorders including depression, anxiety disorders, post-traumatic stress disorder and psychotic disorders. The comparison group was no mental disorders. We only included studies that assessed mental disorders in all TB patients based on the Diagnostic and Statistical Manual of Mental Disorders (DSM), International Classification of Diseases (ICD), or using a validated psychiatric screening instrument. We excluded substance use disorders.

Our primary outcome of interest was TB treatment outcomes as defined by the World Health Organization (WHO): successful (cured, treatment completion) and poor (default or loss to follow-up, failure, death).^11^ Secondary outcomes were diagnostic delay, defined by time from symptom onset to TB diagnosis or treatment initiation; and medication non-adherence measured by self-report, missed visits, pill count, or physiological tests.

#### Data analysis

Records found in database searching were uploaded and de-duplicated in EndNote using the Bramer method.^12^ In duplicate, two reviewers (GL, JS) independently screened abstracts and reviewed the full texts of relevant studies for inclusion. We used Rayyan for screening.^13^

Data from all studies were extracted independently and duplicated by three authors (GL, JS, EM) using a form based on the Cochrane Data Extraction and Assessment template. Data included study characteristics (year, country, study design, sample size) and patient characteristics (type of TB patient, mental health assessment and prevalence, treatment for mental disorder, TB treatment outcomes). Adjusted and unadjusted effect measures (odds, risk, prevalence and hazard ratios) were extracted or calculated for the association between mental disorders and each outcome.

Quality of each study was assessed independently and duplicated among five assessors (GL, JS, EM, JG, AS) using the Risk Of Bias In Non-Randomized Studies of Interventions tool (ROBINS-I).^14^ Based on the literature, we specified age, sex, and socioeconomic status as key *a priori* confounders. If studies adjusted for all three confounders, the best bias judgement attributed for the confounding domain was moderate, according to the ROBINS-I tool.^14^ Disagreements arising in the data extraction and quality assessment process were resolved by consensus.

### Statistical analysis

We calculated odds ratios (OR) and 95% confidence intervals (CI) for the association between mental disorders and treatment outcomes. When direct calculation was not possible, we calculated ORs from risk, hazard and prevalence ratios.^15,16^ Poor outcomes were considered the inverse of successful outcomes. We estimated pooled odds ratios based on the following TB treatment outcomes: any poor outcome (combinations of failure, loss to follow-up, or death), loss to follow-up, and “non-adherence”, which included all definitions of non-adherence across studies. Random-effects models were used, given the clinical heterogeneity of study populations and mental disorders assessed across studies. Heterogeneity was measured using I^2^ statistics.^17^ Analysis was done in R (Vienna, Austria).

## RESULTS

We considered 5,086 studies for inclusion in the review (Figure 1). Abstract screening eliminated 4,931 articles, leaving 155 for full-text screening. An additional 41 articles were screened from citations within included studies. The most common reasons for exclusion were: lack of relevant exposure or outcome (46.2%), outcome not stratified by exposure (28.5%), or mental illness not systematically assessed (7.0%). In total, ten studies met our inclusion criteria, and nine were included in meta-analysis (Figure 1).

**Figure 1.**
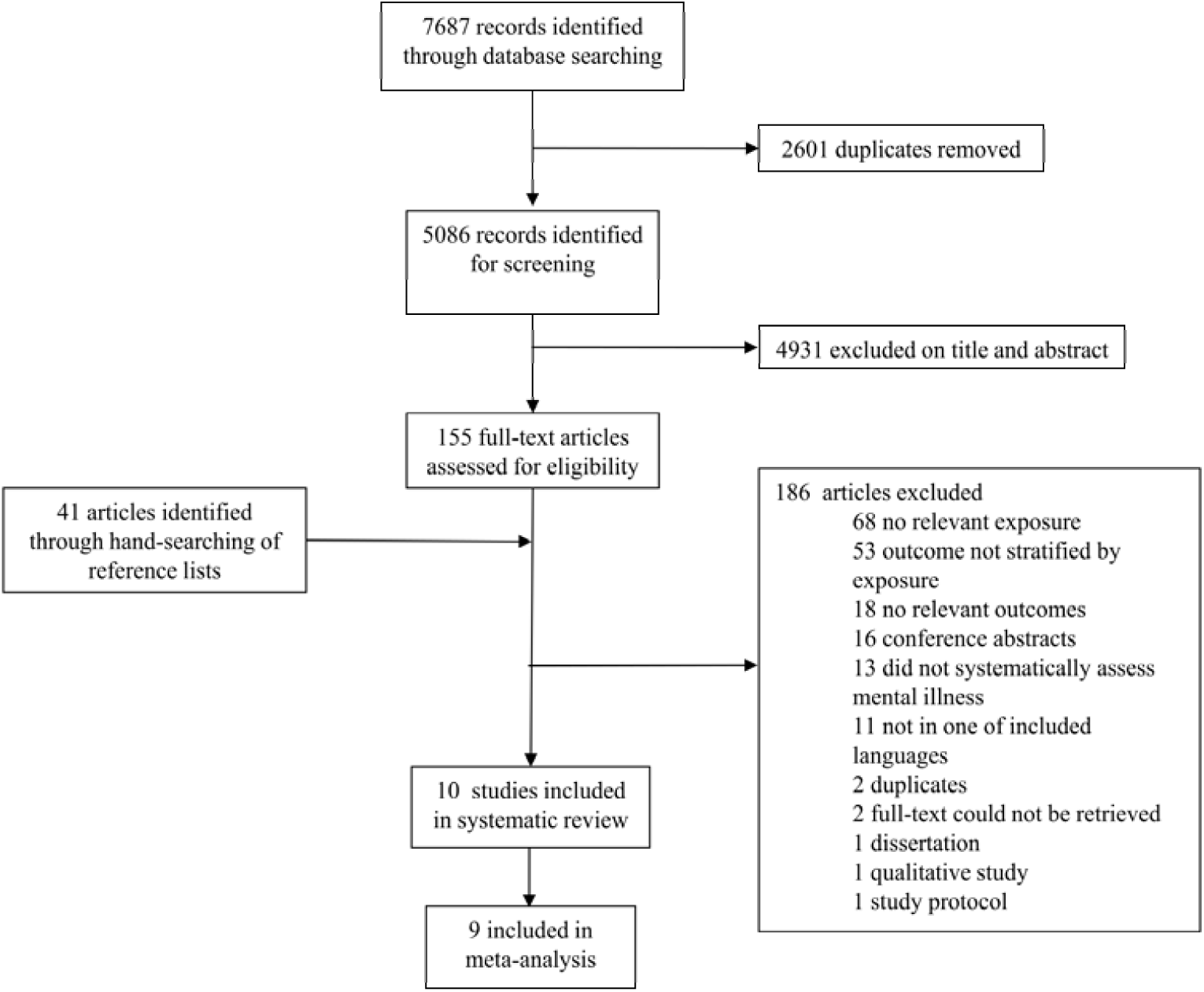
Identification, screening, eligibility, and inclusion of studies, reported using the PRISMA guidelines

### We found no studies on TB treatment delay

#### Characteristics of included studies

The ten included studies reported data on patients beginning TB treatment between 1999 and 2016 (Table 2). They were conducted in sub-Saharan Africa (South Africa, Ethiopia, Zimbabwe, Zambia, and Tanzania) (n=6),^18–23^ Peru (n=3);^24–26^ and China (n=1).^27^ Six studies were cohort studies,^18,19,23–26^ three were cross-sectional studies,^21,22,27^ and one was a case-control study.^20^ The total sample size was 12,868 (range: 49-4,900). Two studies included MDR-TB patients only.^25,26^

**Table 2.**
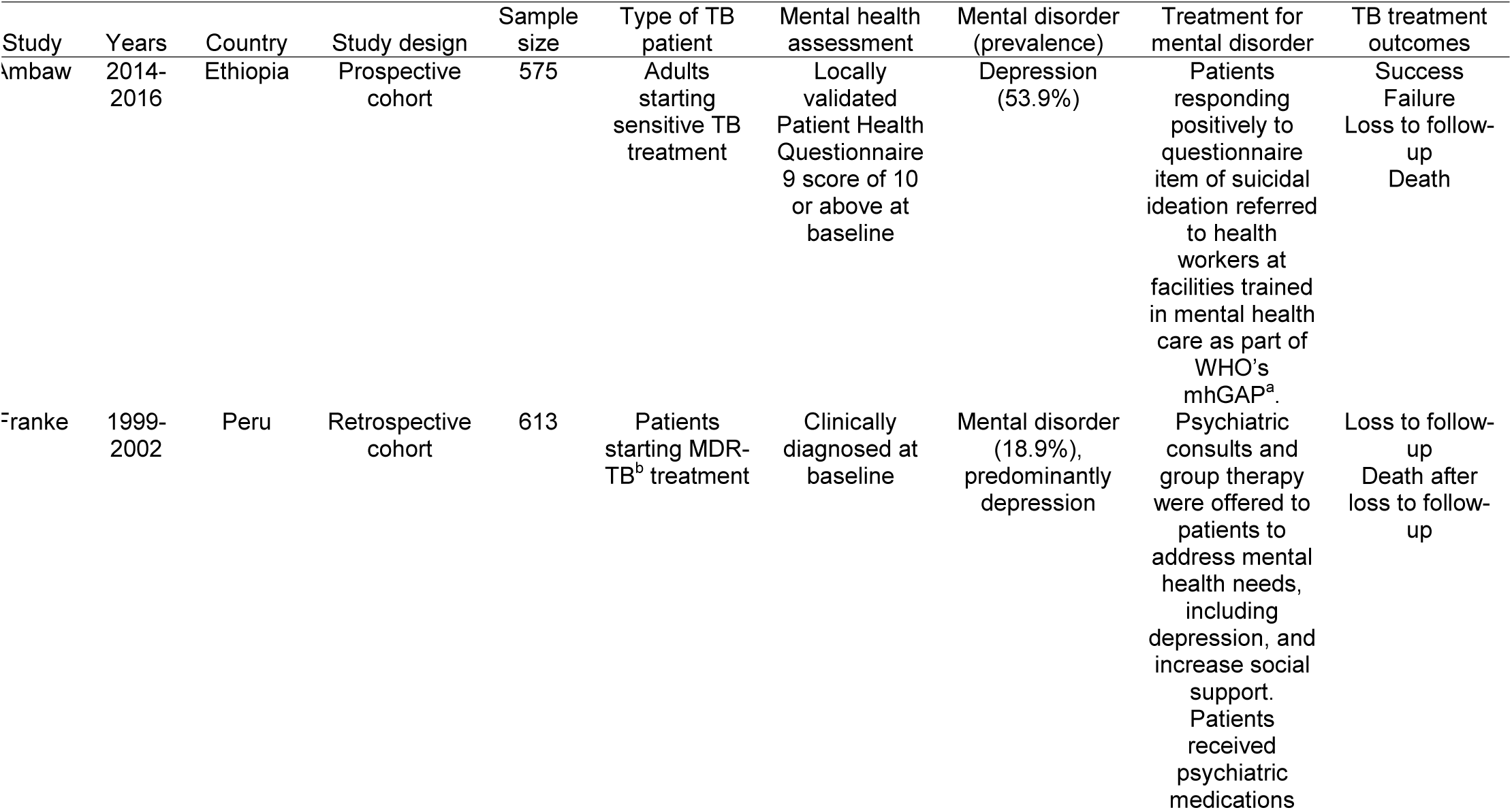
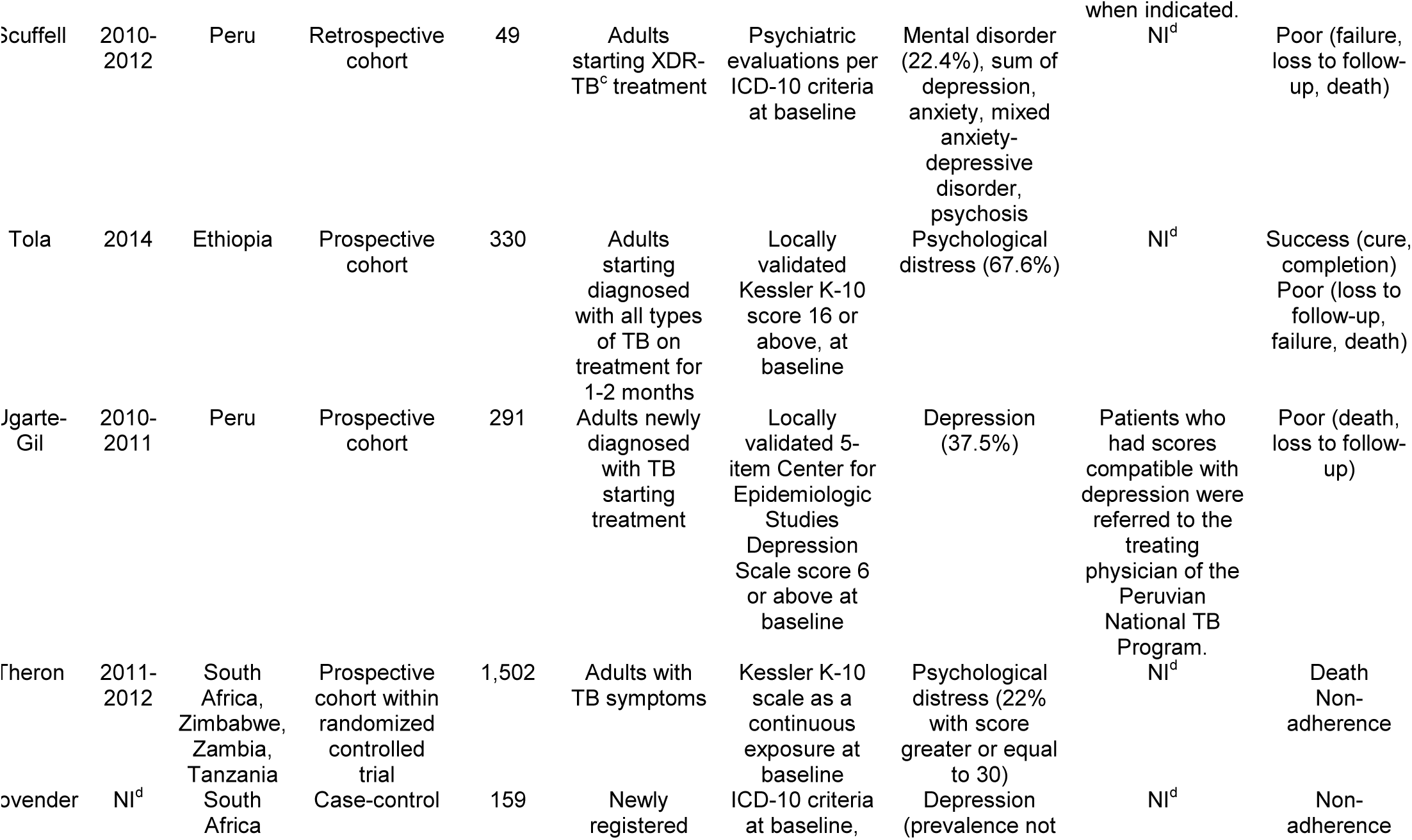
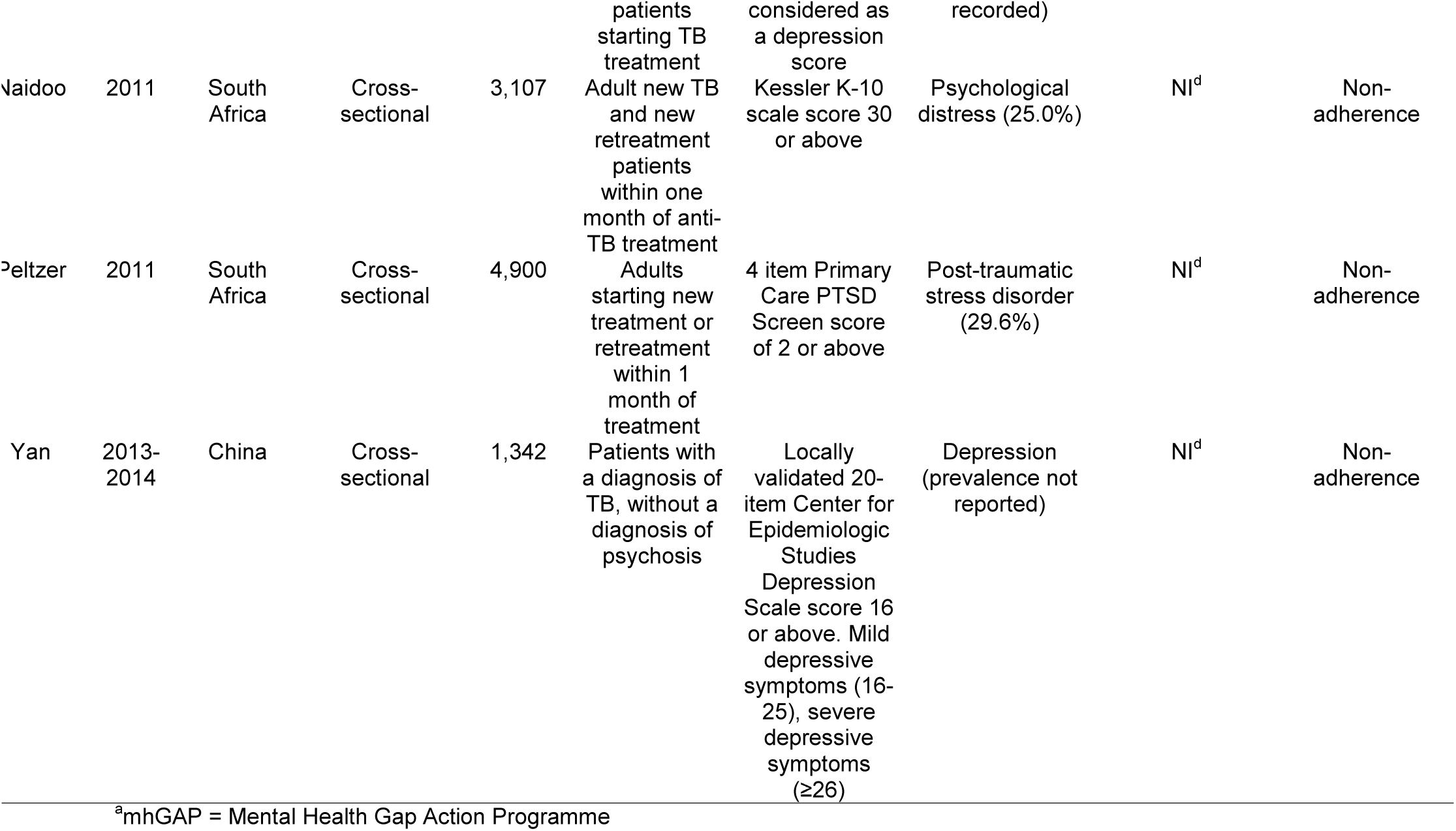
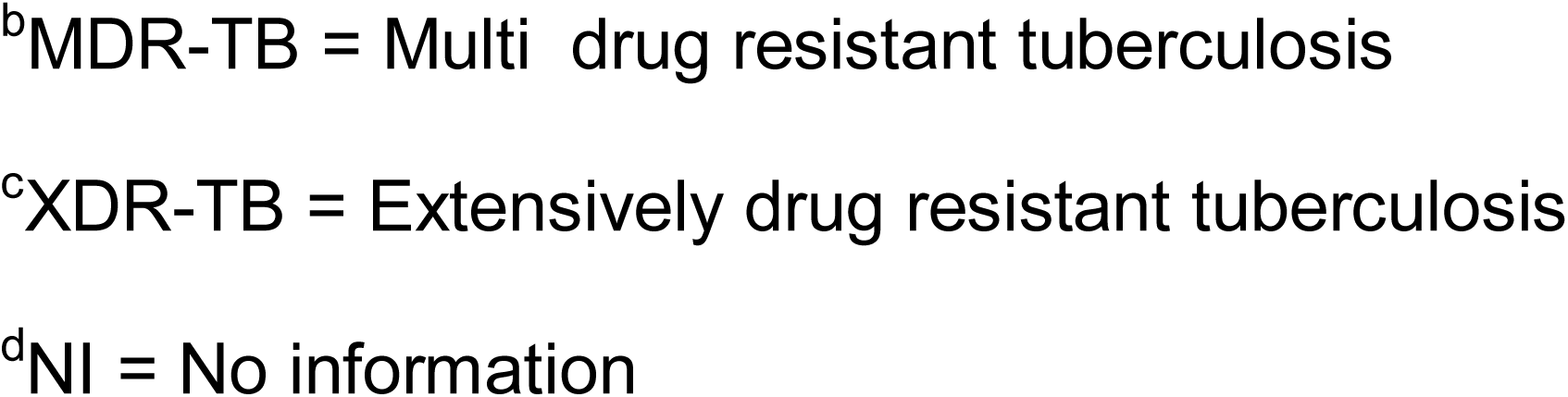
Characteristics of included studies reporting an association between mental disorders and tuberculosis treatment outcomes. SES = socioeconomic status.

Mental disorder was measured at baseline either by psychiatric assessment^20,25,26^ or self-reported screening tools.^18,19,21–24,27^ (Table 2) The prevalence of mental disorders among TB patients ranged from 18.9%^26^ to 67.6%.^19^ Four studies measured depression,^18,20,24,27^ three psychological distress,^19,22,23^ and one post-traumatic stress disorder.^21^ Two studies reported mental disorder as a composite variable.^25,26^ Only two studies indicated that mental health support was provided to patients outside of usual care.^24,26^

### Primary outcomes

#### Association between mental disorders and poor TB treatment outcomes

Four studies estimated the association between mental disorders and poor TB treatment outcomes as a composite measure combining some or all of: treatment failure, loss to follow-up, death (Table 3, Figure 2).^18,19,24,25^ Overall, mental disorders were associated with increased odds of poor TB treatment outcomes (pooled OR 2.13, 95% CI 0.85-5.37, Figure 2). Heterogeneity was high across studies (I^2^ 82%).

**Table 3.**
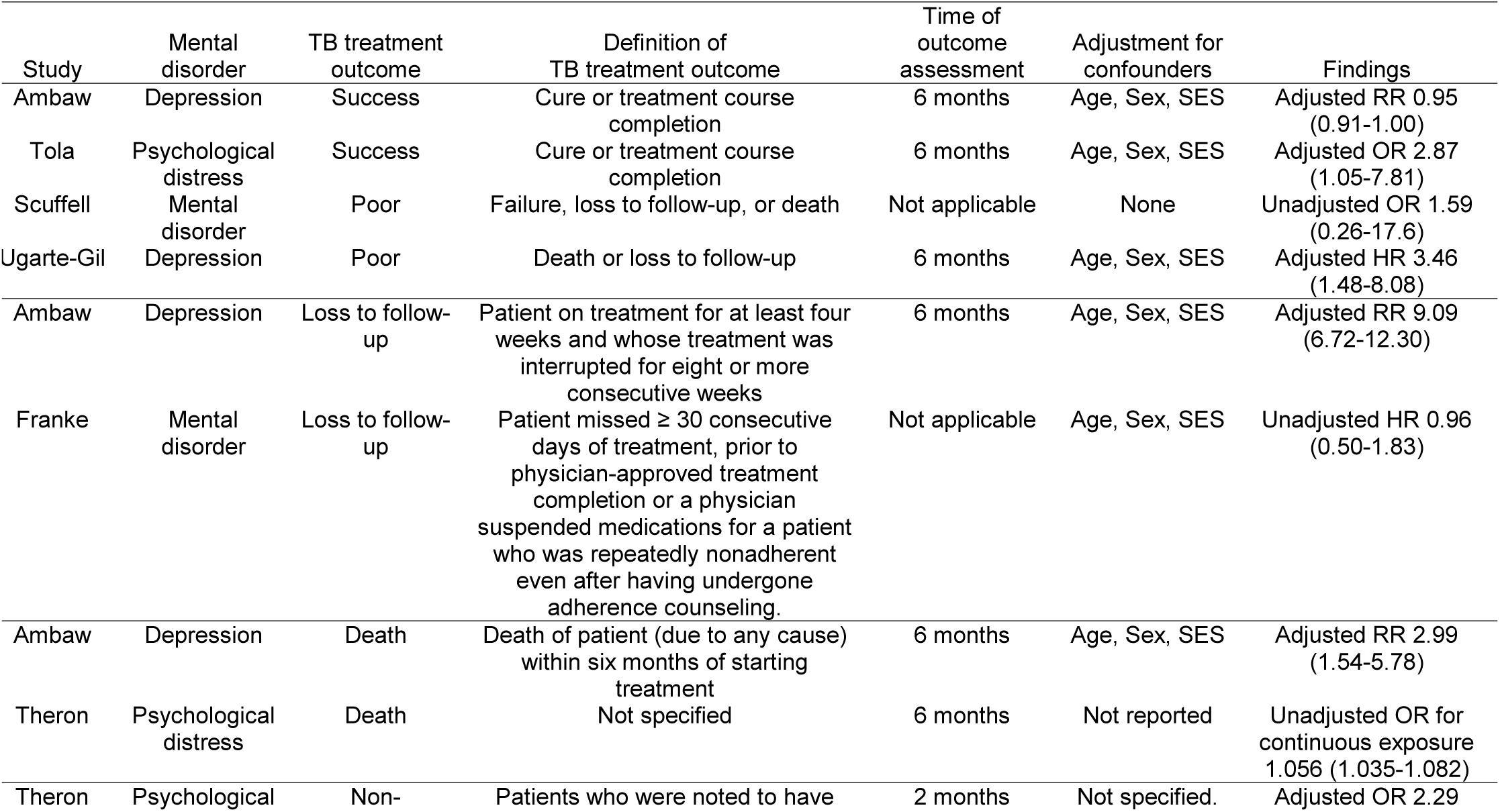
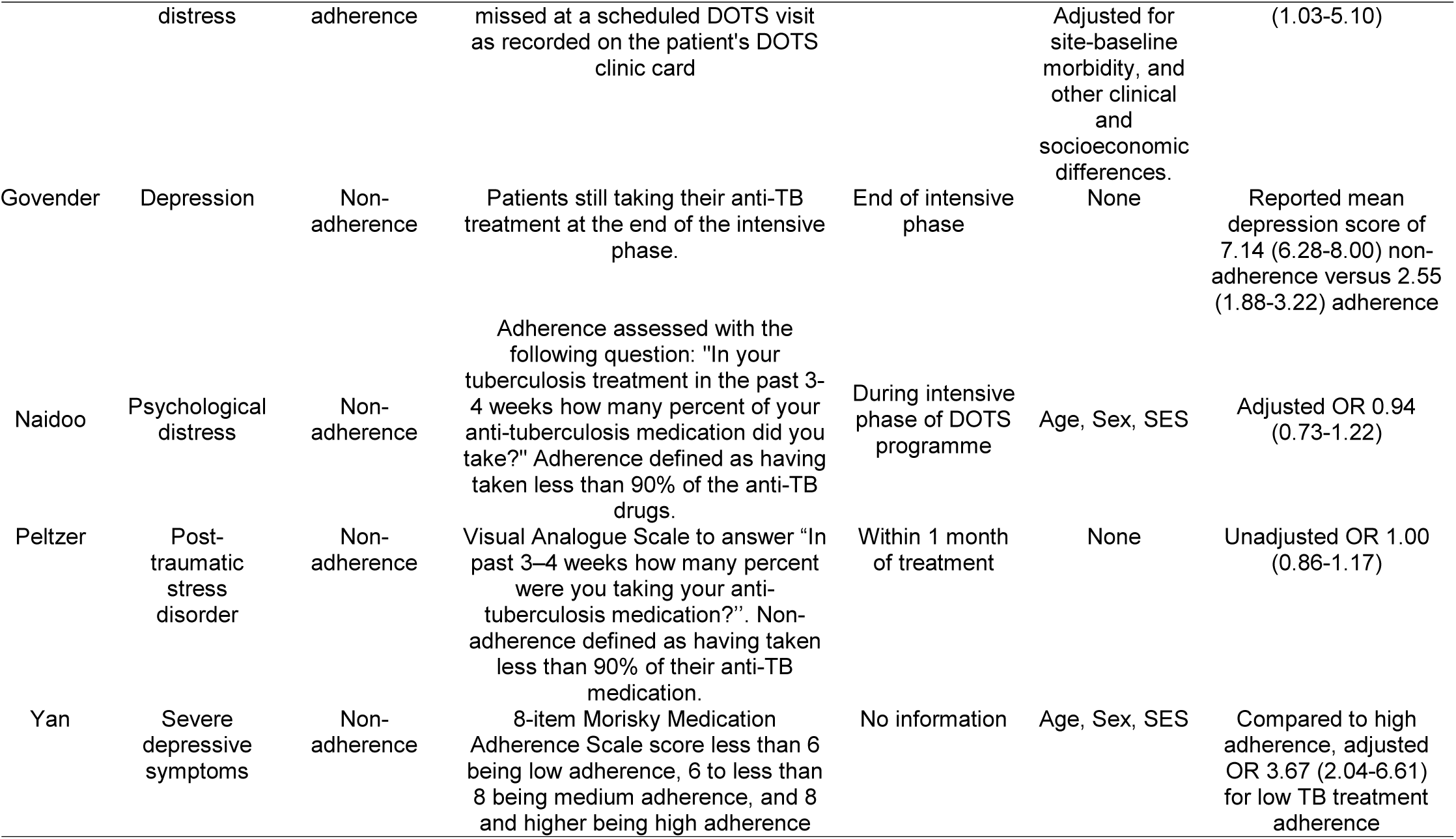
Findings of included studies reporting an association between mental disorders and tuberculosis treatment outcomes

**Figure 2.**
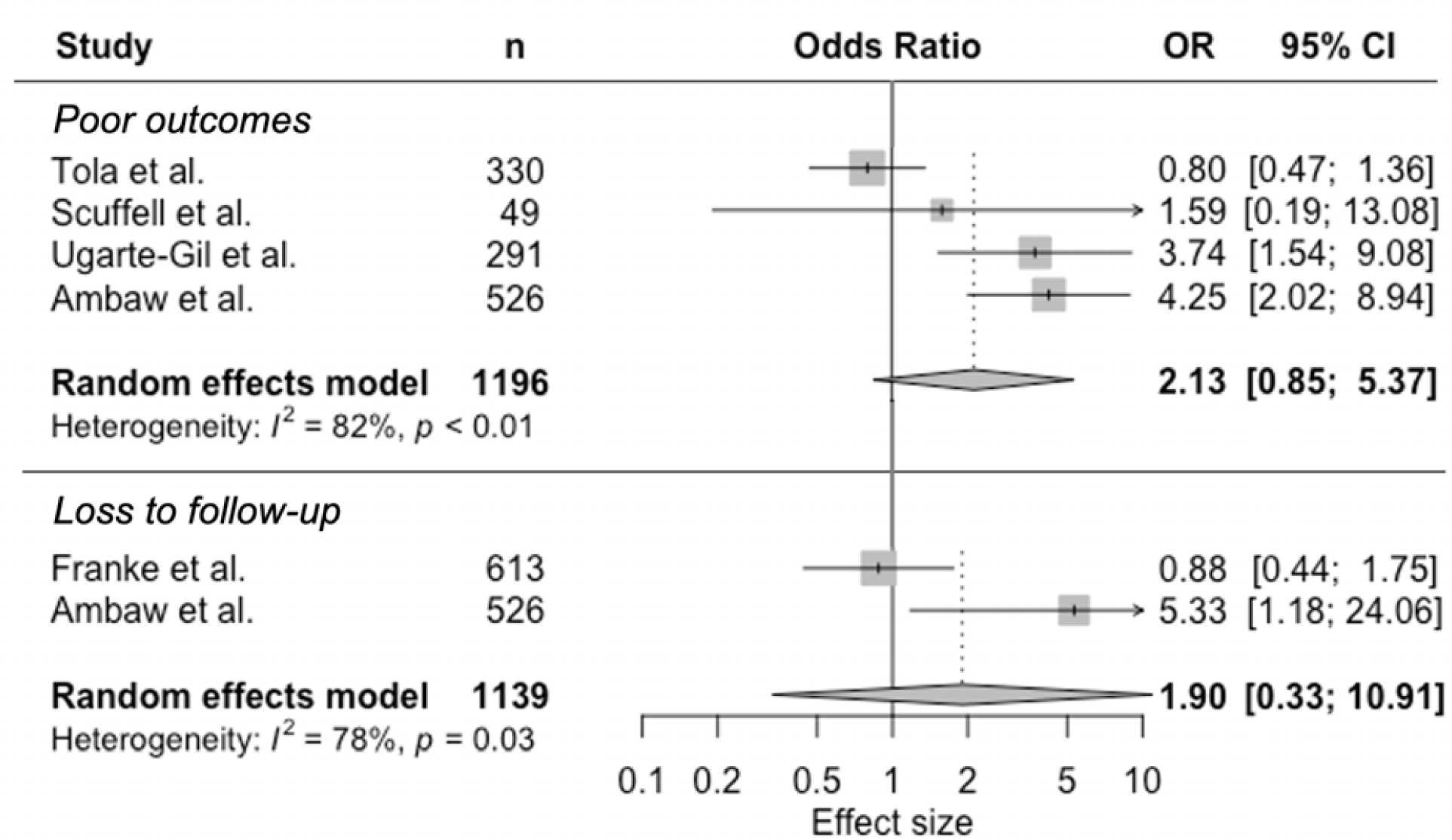
Random-effects meta-analysis of observational studies on mental disorders and tuberculosis treatment outcomes

#### Association between mental disorders and TB treatment loss to follow-up

Two studies focused on the association between mental disorders and loss to follow-up, but used different definitions (Table 3, Figure 2).^18,26^ One study defined loss to follow-up as a patient on treatment for at least four weeks and whose treatment was interrupted for eight or more consecutive weeks.^18^ The other study was defined loss to follow-up as any patient missing 30 or more consecutive days of treatment.^26^ In the meta-analysis (Figure 2), mental disorder was associated with nearly double the odds of loss to follow-up compared with no mental disorder (pooled OR 1.90, 95% CI 0.33-10.91). There was high heterogeneity (I^2^ 78%).

#### Association between mental disorders and death

Two studies reported an association between mental disorders and death (Table 3).^18,23^ In one study, depression was associated with 2.99 times the risk of death (95% CI 1.54-5.78).^18^ Another study revealed that for each point increase of psychological distress score, odds of death increased by 6% (95% CI 4-8%).^23^ Meta-analysis was not conducted for this outcome.

### Secondary outcomes

#### Association between mental disorders and non-adherence to TB treatment

TB treatment non-adherence was examined in five studies, but definitions varied (Table 3, Figure 3). Three studies used self-reported non-adherence,^21,22,27^ one used “discontinuation of treatment beyond the intensive phase of therapy,”^20^ and the fifth defined it as “missing a scheduled DOTS visit”.^23^ One study had insufficient information to calculate an effect estimate and was excluded from the meta-analysis.^20^ Summarizing the effect estimates, there was a trend towards increased odds of non-adherence (OR 1.60, 95% CI 0.84-3.02), with high statistical heterogeneity (I^2^ 86%) (Figure 3).

**Figure 3.**
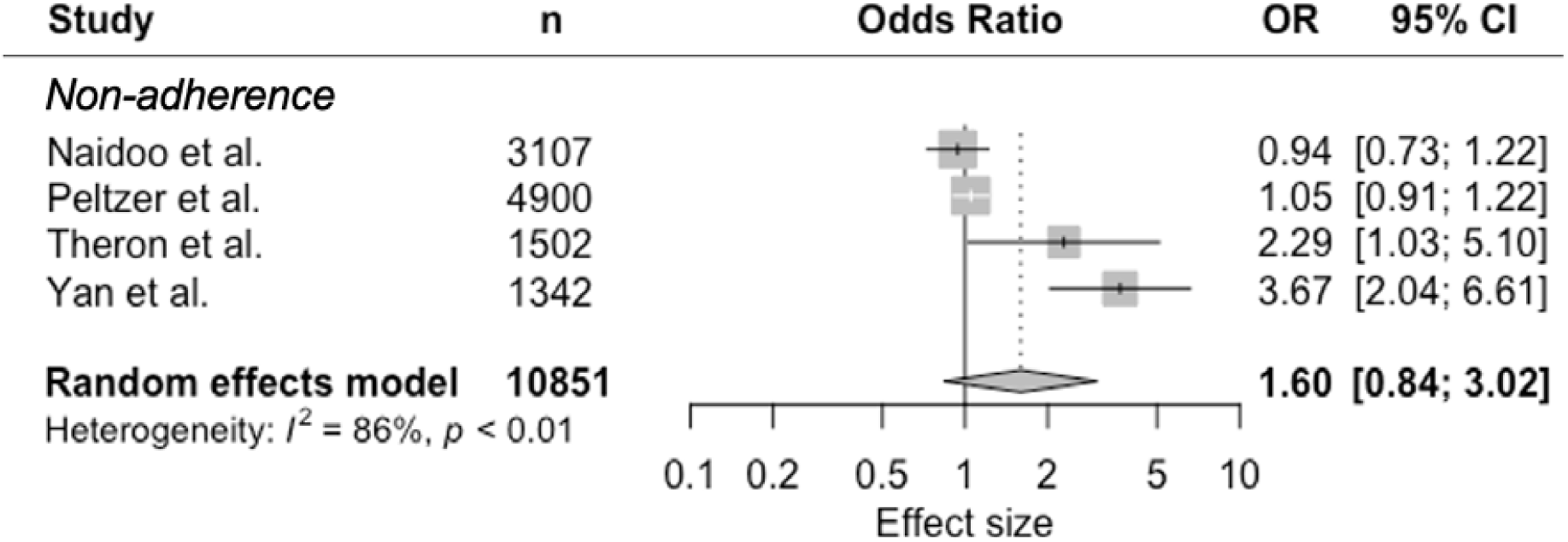
Random-effects meta-analysis of observational studies on mental disorders and tuberculosis treatment non-adherence

### Risk of bias assessment

All included studies were observational studies, putting them at risk of bias according to ROBINS-I (Table 4). Half of the studies showed moderate risk of bias related to confounders by adjusting results for age, sex and socioeconomic status,^18,22–24,27^ while the other half displayed serious risk of bias by not accounting for these confounders.^19– 21,25,26^ Five studies had low risk of bias in selection of participants,^18,23–26^ but the other five studies were at serious risk of bias with study inclusion being related to mental health status and TB treatment outcome.^19–22,27^ The most serious methodological concern was risk of classification of exposure. Six studies were at serious risk of bias for this domain, either because mental health was determined at the same time as the outcome or because the definition of mental disorder was not clear.^19–23,27^ The other four studies had low risk of bias because they used locally validated tools or psychiatric interviews.^18,24–26^ Two studies reported the provision of psychiatric treatment outside of usual care and were classified as serious risk of bias for deviations from intended exposure.^24,26^ The other studies were either at low risk of bias or had insufficient information.^18–23,25,27^ For missing data one study was at serious risk of bias,^20^ seven were at low risk of bias,^18,19,22,23,25–27^ and two had no information.^21,24^ Four studies used self-reported measures and were classified as serious risk of bias due to measurement of outcomes.^20–22,27^ All except one study^22^ presented a low risk of bias in selection of reported results.

**Table 4.**
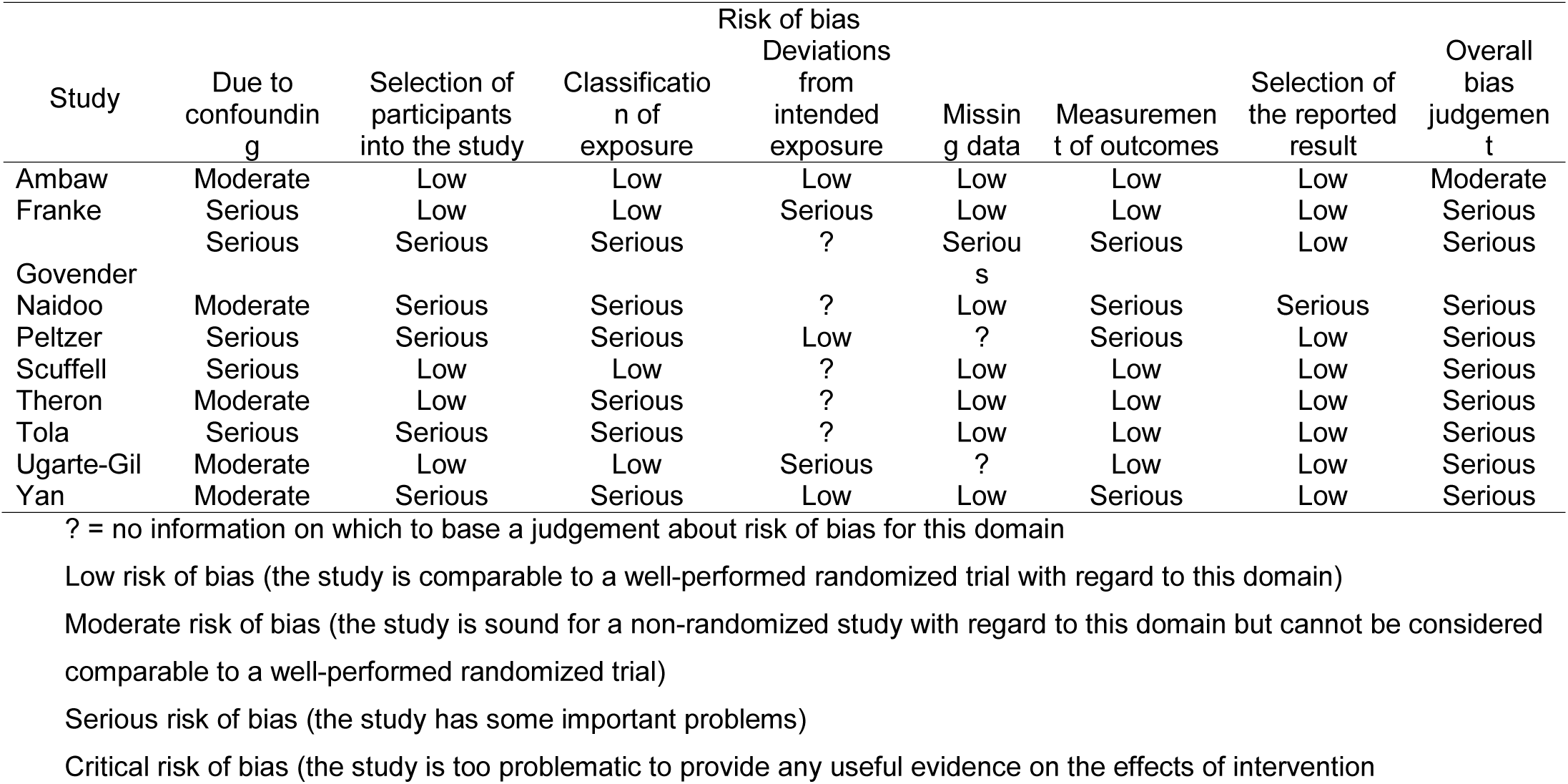
Risk of bias assessment of observational studies on mental disorder and TB treatment outcomes and non-adherence, according to ROBINS-I criteria

## DISCUSSION

Our systematic review and meta-analysis found TB patients with mental disorders may have double the risk of loss to follow-up and three times the risk of death compared with patients without mental disorders. Mental disorders were also associated with twice the risk of treatment non-adherence. Still, few studies met criteria for inclusion, resulting in decreased precision of effect estimates, and most included studies had serious risk of bias.

These results are similar to but weaker than associations reported in another recent systematic review focused only on depression.^28^ This difference may be partially due to variations in outcome definitions and statistical approaches between studies. The strong link between mental disorders and non-adherence to TB treatment is consistent with studies examining medication adherence in patients with mental disorders comorbid with HIV and non-communicable diseases.^29–31^

We found highly heterogeneous definitions of mental disorders across studies. Of the ten included studies, only three captured mental disorders using ICD or DSM criteria.^20,25,26^ The remaining seven used screening tools with various cut-off scores.^18,19,21–24,27^ Given the overlap between some symptoms of TB and mental disorders (e.g. disturbances in appetite, energy, and sleep), screening tools may not be as effective at distinguishing etiology.^32^ Although half of the studies using screening tools described validation in local populations, none described specific validation among TB patients. This validation is imperative to better evaluate mental health in this population.^33^

There was also variation in definitions of TB outcomes. The WHO defines loss to follow-up as “a TB patient who did not start treatment or whose treatment was interrupted for 2 consecutive months or more.”^11^ However, only one study used this definition;^19^ the other defined loss to follow-up as one month of treatment interruption.^26^ The five studies evaluating TB treatment adherence^20–23,27^ had even more disparate definitions: yes or no,^20^ taking less than 90% of medication,^21,22^ missing a scheduled DOTS visit,^23^ and a score qualifying low, medium, or high adherence.^27^ Variations in outcome definitions may partially explain differences in magnitude and precision of these results.

All included studies except one^18^ had serious risk of bias using the ROBINS-I tool,^18^ which limits the interpretation of current evidence. Half of the included studies did not adjust for important confounders of age, sex, and socioeconomic status,^32^ and we could not account for these in meta-analysis. Residual confounding is likely present since none of the studies measured and adjusted for malnutrition, substance, alcohol or tobacco use.^34–37^ There are also likely variations in quality and expectations of TB care between studies.

Only one study specifically described mental health intervention provided.^26^ MDR-TB patients in Peru received group therapy and psychiatric medications to facilitate adherence, which may explain why the association between mental disorders and loss to follow-up was not significant in the study.^26^ Interventions that aim to directly support TB patients (e.g. psychological therapies and anti-depressant treatment) have been shown to be acceptable^38,39^ and improve TB outcomes.^40^ Individual psychotherapy, peer-led “TB clubs”, and psychotropic medications have been shown to improve adherence to TB treatment and TB.^10,41^ Patient-centered approaches like these either address mental health directly or address TB-related stigma, but are not widely adopted.

### Strengths and limitations of study

Our meta-analysis was limited by the small number of studies that met inclusion criteria. For example, 13 studies from retrospective chart review or registry data were excluded because it was unclear if mental disorders were assessed for all patients. To mitigate the small number of studies, we combined all types of mental disorders and both TB and MDR-TB together in the meta-analyses, resulting in high levels of heterogeneity and wide confidence intervals in our pooled estimates. For example, compared to drug-sensitive TB, psychosocial stressors associated with MDR-TB are significantly more complicated;^33,42^ treatment regimens often include medicines with psychiatric side effects, and treatment duration is much longer.^43^ As a result, we were unable to form conclusions for specific mental disorders, although different mental disorders may have different impacts on TB outcomes.

## CONCLUSIONS

This review emphasizes the scarcity of studies that methodically evaluate the impact of mental disorders on TB treatment outcomes. A strong evidence base is essential to close the treatment gap in mental health care and accelerate progress towards achieving the targets of the WHO’s End-TB Strategy. Future studies should systematically assess mental disorders in TB patients using locally validated screening tools or psychiatric evaluations, use standard outcome definitions, and measure and control for confounders. Improving the quality of future studies will showcase the magnitude of associations between mental disorders and TB treatment outcomes.

## Data Availability

Data used in the manuscript is available on request to the authors.

## ACKNOWLEDGEMENTS

We thank Paul Mason, Fred Andayi, Lindokuhle Ndlandla, Lekha Puri, Ajaj Rangaraj, and Rafael Silva Duarte for proposing this systematic review and meta-analyses during a small group session at the McGill Summer Institute of Infectious Diseases.

Funding was provided in part by the US National Institute of Mental Health (NIMH grant K01 MH104514) and National Institute of Allergy and Infectious Diseases (NIAID grant K01 AI118559).

## REFERENCES

1. World Health Organization. Implementing the end TB strategy: the essentials. Geneva, Switzerland: World Health Organization; 2015.

2. Doherty AM, Kelly J, McDonald C, O’Dywer AM, Keane J, Cooney J. A review of the interplay between tuberculosis and mental health. Gen Hosp Psychiatry. 2013;35(4):398–406.

3. Ribeiro Macedo L, Reis-Santos B, Riley LW, Maciel EL. Treatment outcomes of tuberculosis patients in Brazilian prisons: a polytomous regression analysis. Int J Tuberc Lung Dis. 2013 Nov;17(11):1427–34.

4. Duarte EC, Bierrenbach AL, Barbosa da Silva J, Tauil PL, de Fátima Duarte E. Factors associated with deaths among pulmonary tuberculosis patients: a case-control study with secondary data. J Epidemiol Community Health. 2009;63(3):233–8.

5. Wang WB, Zhao Q, Yuan ZA, Jiang WL, Liu ML, Xu B. Deaths of tuberculosis patients in urban China: a retrospective cohort study. Int J Tuberc Lung Dis. 2013 Apr;17(4):493–8.

6. Garrido M da S, Penna ML, Perez-Porcuna TM, de Souza AB, Marreiro L da S, Albuquerque BC, et al. Factors associated with tuberculosis treatment default in an endemic area of the Brazilian Amazon: a case control-study. PLoS One. 2012 Jun 12;7(6):e39134.

7. Sweetland AC, Jaramillo E, Wainberg ML, Chowdhary N, Oquendo MA, Medina-Marino A, et al. Tuberculosis: an opportunity to integrate mental health services in primary care in low-resource settings. Lancet Psychiatry. 2018 Dec;5(12):952–4.

8. Kuo S-C, Chen Y-T, Li S-Y, Lee Y-T, Yang AC, Chen T-L, et al. Incidence and outcome of newly-diagnosed tuberculosis in schizophrenics: a 12-year, nationwide, retrospective longitudinal study. BMC Infect Dis. 2013 Jul 29;13:351.

9. Sikjær MG, Løkke A, Hilberg O. The influence of psychiatric disorders on the course of lung cancer, chronic obstructive pulmonary disease and tuberculosis. Respir Med. 2018;135:35–41.

10. Sweetland A, Oquendo M, Wickramaratne P, Weissman M, Wainberg M. Depression: a silent driver of the global tuberculosis epidemic. World Psychiatry. 2014 Oct;13(3):325–6.

11. World Health Organization. Definitions and reporting framework for tuberculosis - 2013 revision. Geneva, Switzerland: World Health Organization; 2013.

12. Bramer WM, Giustini D, de Jonge GB, Holland L, Bekhuis T. De-duplication of database search results for systematic reviews in EndNote. J Med Libr Assoc. 2016 Jul;104(3):240–3.

13. Ouzzani M, Hammady H, Fedorowicz Z, Elmagarmid A. Rayyan—a web and mobile app for systematic reviews. Syst Rev. 2016;5(1):210.

14. Sterne JA, Hernán MA, Reeves BC, Savović J, Berkman ND, Viswanathan M, et al. ROBINS-I: a tool for assessing risk of bias in non-randomised studies of interventions. BMJ. 2016 Oct 12;355:i4919.

15. Zhang J, Yu KF. What’s the relative risk? A method of correcting the odds ratio in cohort studies of common outcomes. JAMA. 1998 Nov 18;280(19):1690–1.

16. Tierney JF, Stewart LA, Ghersi D, Burdett S, Sydes MR. Practical methods for incorporating summary time-to-event data into meta-analysis. Trials. 2007;8:16.

17. Higgins JPT, Thompson SG, Deeks JJ, Altman DG. Measuring inconsistency in meta-analyses. BMJ. 2003;327(7414):557–60.

18. Ambaw F, Mayston R, Hanlon C, Medhin G, Alem A. Untreated depression and tuberculosis treatment outcomes, quality of life and disability, Ethiopia. Bull World Health Organ. 2018;96(4):243–55.

19. Tola HH, Shojaeizadeh D, Garmaroudi G, Tol A, Yekaninejad MS, Ejeta LT, et al. Psychological distress and its effect on tuberculosis treatment outcomes in Ethiopia. Glob Health Action. 2015;8:29019.

20. Govender S, Mash R. What are the reasons for patients not adhering to their anti-TB treatment in a South African district hospital? AJOL [Internet]. 2009 [cited 2020 Mar 9];51(6). Available from: https://www.ajol.info/index.php/safp/article/view/50083

21. Peltzer K, Naidoo P, Matseke G, Louw J, McHunu G, Tutshana B. Prevalence of post-traumatic stress symptoms and associated factors in tuberculosis (TB), TB retreatment and/or TB-HIV co-infected primary public health-care patients in three districts in South Africa. Psychol Health Med. 2013;18(4):387–97.

22. Naidoo P, Peltzer K, Louw J, Matseke G, McHunu G, Tutshana B. Predictors of tuberculosis (TB) and antiretroviral (ARV) medication non-adherence in public primary care patients in South Africa: a cross sectional study. BMC Public Health. 2013 Apr 26;13:396.

23. Theron G, Peter J, Zijenah L, Chanda D, Mangu C, Clowes P, et al. Psychological distress and its relationship with non-adherence to TB treatment: a multicentre study. BMC Infect Dis. 2015;15(1):253.

24. Ugarte-Gil C, Ruiz P, Zamudio C, Canaza L, Otero L, Kruger H, et al. Association of major depressive episode with negative outcomes of tuberculosis treatment. PLoS One. 2013;8(7):e69514.

25. Scuffell J, Boccia D, Garcia Velarde F, Leon SR, Raviola G, Lecca L, et al. Mental disorders and drug/alcohol use in patients commencing extensively drug-resistant tuberculosis treatment. Public Health Action. 2017 Sep 21;7(3):237–9.

26. Franke MF, Appleton SC, Bayona J, Arteaga F, Palacios E, Llaro K, et al. Risk factors and mortality associated with default from multidrug-resistant tuberculosis treatment. Clin Infect Dis. 2008;46(12):1844–51.

27. Yan S, Zhang S, Tong Y, Yin X, Lu Z, Gong Y. Nonadherence to antituberculosis medications: the impact of stigma and depressive symptoms. Am J Trop Med Hyg. 2018 Jan;98(1):262–5.

28. Ruiz-Grosso P, Cachay R, Flor A de la, Schwalb A, Ugarte-Gil C. Association between tuberculosis and depression on negative outcomes of tuberculosis treatment: a systematic review and meta-analysis. PLoS One. 2020 Jan 10;15(1):e0227472.

29. Wang PS, Bohn RL, Knight E, Glynn RJ, Mogun H, Avorn J. Noncompliance with antihypertensive medications: the impact of depressive symptoms and psychosocial factors. J Gen Intern Med. 2002 Jul;17(7):504–11.

30. Eze-Nliam CM, Thombs BD, Lima BB, Smith CG, Ziegelstein RC. The association of depression with adherence to antihypertensive medications: a systematic review. J Hypertens. 2010;28(9):1785–95.

31. Uthman OA, Magidson JF, Safren SA, Nachega JB. Depression and adherence to antiretroviral therapy in low-, middle- and high-income countries: a systematic review and meta-analysis. Curr HIV/AIDS Rep. 2014 Sep;11(3):291–307.

32. Sweetland AC, Kritski A, Oquendo MA, Sublette ME, Norcini Pala A, Silva LRB, et al. Addressing the tuberculosis-depression syndemic to end the tuberculosis epidemic. Int J Tuberc Lung Dis. 2017 01;21(8):852–61.

33. Walker IF, Baral SC, Wei X, Huque R, Khan A, Walley J, et al. Multidrug-resistant tuberculosis treatment programmes insufficiently consider comorbid mental disorders. Int J Tuberc Lung Dis. 2017;21(6):603–9.

34. Deiss RG, Rodwell TC, Garfein RS. Tuberculosis and illicit drug use: review and update. Clin Infect Dis. 2009 Jan 1;48(1):72–82.

35. Janse Van Rensburg A, Dube A, Curran R, Ambaw F, Murdoch J, Bachmann M, et al. Comorbidities between tuberculosis and common mental disorders: a scoping review of epidemiological patterns and person-centred care interventions from low-to-middle income and BRICS countries. Infect Dis Poverty. 2020;9(1):4.

36. Balinda IG, Sugrue DD, Ivers LC. More than malnutrition: a review of the relationship between food insecurity and tuberculosis. Open Forum Infect Dis. 2019 Mar 7;6(4):ofz102.

37. Najt P, Fusar-Poli P, Brambilla P. Co-occurring mental and substance abuse disorders: a review on the potential predictors and clinical outcomes. Psychiatry Res. 2011 Apr 30;186(2):159–64.

38. Walker IF, Khanal S, Hicks JP, Lamichhane B, Thapa A, Elsey H, et al. Implementation of a psychosocial support package for people receiving treatment for multidrug-resistant tuberculosis in Nepal: a feasibility and acceptability study. PLoS One. 2018 Jul 26;13(7):e0201163.

39. Wingfield T, Tovar MA, Huff D, Boccia D, Montoya R, Ramos E, et al. A randomized controlled study of socioeconomic support to enhance tuberculosis prevention and treatment, Peru. Bull World Health Organ. 2017 Apr 1;95(4):270–80.

40. Rocha C, Montoya R, Zevallos K, Curatola A, Ynga W, Franco J, et al. The innovative socio-economic interventions against tuberculosis (ISIAT) project: an operational assessment. Int J Tuberc Lung Dis. 2011 Jun;15(Suppl 2):S50–7.

41. Alipanah N, Jarlsberg L, Miller C, Linh NN, Falzon D, Jaramillo E, et al. Adherence interventions and outcomes of tuberculosis treatment: a systematic review and meta-analysis of trials and observational studies. PLoS Med. 2018;15(7):e1002595.

42. Tanimura T, Jaramillo E, Weil D, Raviglione M, Lönnroth K. Financial burden for tuberculosis patients in low- and middle-income countries: a systematic review. Eur Respir J. 2014 Jun 1;43(6):1763–75.

43. Khanal S, Elsey H, King R, Baral SC, Bhatta BR, Newell JN. Development of a patient-centred, psychosocial support intervention for multi-drug-resistant tuberculosis (MDR-TB) care in Nepal. PLoS One. 2017;12(1):e0167559.

